# Unscheduled bleeding and endometrial cancer in women on postmenopausal hormone replacement therapy and their matched controls: protocol for a descriptive cohort study using the Orchid-e database

**DOI:** 10.64898/2026.04.17.26350707

**Authors:** Margaret Smith, Sharon Dixon, Sheba Ziyenga, Jennifer Hirst, Clare Bankhead, Brian Nicholson

## Abstract

Hormone replacement therapy (HRT) with oestrogen and progestogen is a common medical treatment for alleviating symptoms of menopause. Since 2015, its use has been increasing in the UK. Unscheduled bleeding can be a symptom of endometrial cancer, and guidelines state that women experiencing this should have an urgent referral for suspected endometrial cancer. However, unscheduled bleeding is also common in women taking HRT, particularly in the first few months after starting HRT or if there is a change in regimen. Current guidelines may result in women on HRT receiving referrals that are not necessary and undergoing unpleasant and invasive tests such as hysteroscopy. However, there is a lack of current information to guide recommendations.

This protocol describes a cohort study in the ORCHID-e database of anonymised patient records from English primary care. We will use a cohort of women aged over 40 years starting on HRT with oestrogen and progestogen, age matched to women who have not started HRT. Exposure will be a prescription for oestrogen containing HRT with no previous prescription for oestrogen containing HRT in the previous year. Index date in each matched set will be the date of this prescription.

Prescriptions for progestogen containing drugs will not be used to define the exposure, but this information will be extracted to describe the study population and for sensitivity analyses. Outcomes will be consultations for unscheduled bleeding, urgent referrals for suspected endometrial cancer, and diagnosis of endometrial cancer. Women will be followed up until they change exposure status or are otherwise censored. Women who start taking HRT in follow-up will re-enter the cohort in the exposed group.

We will describe proportions of women with a code for consulting with unscheduled bleeding, proportions of those women referred for further investigation on the pathway for suspected endometrial cancer, and proportions diagnosed with endometrial cancer within one year of referral. We will investigate the diagnostic accuracy of unscheduled bleeding for endometrial cancer separately for women on HRT and those not on HRT. Analyses will be done by 6-month categories of time since index, age, calendar year, sociodemographic variables, risk factors for endometrial cancer, type of HRT.

**Lay Summary:** Menopause is when a woman’s periods stop for good. This usually happens between the ages of 45 and 55, but it can happen earlier. During menopause, the level of the hormone oestrogen falls, which can cause symptoms such as hot flushes, night sweats, poor sleep, low mood and vaginal dryness. For some women these symptoms are mild, but for others they can be very difficult and affect daily life. Many women use hormone replacement therapy (HRT) to help manage symptoms of the menopause by replacing some of the hormones their body no longer makes. The use of HRT has been increasing, including higher doses, starting at younger and finishing at older ages.

Bleeding is a common side effect in women starting on HRT. It is also a symptom of womb cancer. Current guidelines for womb cancer are that women who have unscheduled bleeding after the menopause should have an urgent referral for further tests for possible cancer. However, these guidelines were written when fewer women were taking HRT. Some women on HRT might be undergoing unnecessary investigations for suspected womb cancer. Right now, doctors do not have enough clear information to know if current guidance needs to change.

In this study, we will use safely stored health records where no one can be identified. We will look at women aged 40 and over who have recently started HRT, and compare them with women of the same age who are not using HRT. By following this information over time, we can see what happens to them.

**We will look at:** How often women and other people with a womb contact their GP because of unscheduled bleeding.

How often they are then sent for urgent tests that check for cancer.

How often cancer is found within a year of these tests.

How useful unscheduled bleeding is as a warning sign of cancer for people using HRT.

If there is enough information, we will also look at whether results are different for people depending on factors such as how long they have been using HRT, their age, the type of HRT they take, their background, where they live, their general health, and their social class.

The results will help us to understand how women on HRT with unscheduled bleeding are referred for further investigations on the suspected cancer pathway. They will provide evidence for the development of new guidelines for unexpected bleeding and cancer of the womb that are more relevant to women on HRT.

## Background

Menopause usually occurs around ages 45-55 years, but some women experience early menopause at around ages 40-44. Premature menopause, before the age of 40 years, can occur naturally or in association with surgery or treatment for other conditions (for example chemotherapy or cancer treatment). Menopause can be relatively asymptomatic or it can be associated with a wide range of symptoms due to a lack of oestrogen, including hot flushes and night sweats, vaginal dryness, difficulty sleeping, low mood. Symptom severity can vary from minor to highly debilitating and can occur over short or long periods. After menopause, there is a higher risk of certain conditions such as osteoporosis or cardiovascular disease.

Hormone replacement therapy (HRT) with oestrogen and progestogen is a common medical treatment for alleviating symptoms of menopause. Historically HRT use has fluctuated against a backdrop of emerging epidemiological literature, which has been widely publicised in the media. Since 2015, its use has been increasing in the UK in a context of advocacy and media attention on menopause [1-3]. In England in 2023/24 there were 13 million HRT items prescribed, a 22% increase from 2022/23 [3]. Nearly 20% of women aged 45-55 were taking HRT in 2024 [4].

Incidence of uterine or womb cancer (which is most commonly endometrial cancer) increases with age from 7.5 cases per 100,000 in women aged 40-44 years in 2017-2019, to 32.9 in those age 50-54 years and 96.9 at age 75-79 years [5]. Oestrogen therapy in HRT increases the risk of endometrial hyperplasia and endometrial cancer. This risk is mitigated by co-administration of progestogens, either cyclically (sequential HRT) or continually (continuous combined HRT)[6]. Combined HRT may provide slightly better endometrial protection than sequential but there is also variation within regimens, for example dependent on duration of use, or oestrogen dose and number of days of progestogen within sequential HRT [6, 7].

Unscheduled bleeding (sometimes called post-menopausal bleeding) is a common symptom of endometrial cancer [8]. Guidelines state that women experiencing unscheduled bleeding should be given an urgent referral for further tests on the suspected gynaecological cancer pathway [9].

Unscheduled bleeding is also common in women taking HRT, particularly in the first few months after starting it or if there is a change in regimen [10]. Therefore, these guidelines may result in women on HRT receiving referrals that are not necessary and undergoing unpleasant and invasive tests such as hysteroscopy. On behalf of The British Menopause Society an expert panel has developed guidelines more appropriate to women on HRT that allow for bleeding pattern, HRT regimen and other endometrial cancer risk factors [10]. However, there is a lack of current information to guide recommendations, including about patterns of unscheduled bleeding in women on HRT, subsequent referrals and endometrial cancer diagnoses. In addition, as HRT use has increasingly been used for control of menopause symptoms, so have the numbers of women taking it at 60 years or older. Higher doses of oestrogen are also sometimes prescribed. The risk of endometrial cancer and its relationship to unscheduled bleeding is even less well understood in these groups.

In this study, we will estimate the incidence of unscheduled bleeding in women on HRT compared to women not on HRT. We will also investigate the incidence of subsequent urgent referrals for suspected endometrial cancer and subsequent diagnoses of endometrial cancer in these women.

We will do this by time since started HRT, age of woman and sociodemographic variables so as to get an accurate idea of the incidence of unscheduled bleeding in different groups and how this leads on to urgent referrals for suspected cancer and subsequent diagnosis of endometrial cancer. We will utilise the Oxford Research Clinical Informatics Hub Epidemiology Platform (ORCHID-e) database, which is an anonymised database of electronic health records (EHR) for epidemiological research with a focus on cancer. The results will help to decide which women with unscheduled bleeding to prioritise for further investigation, and to clarify the risk of endometrial cancer in different subgroups.

### Patient and Public Involvement and Engagement (PPIE)

The protocol summary was reviewed by a PPIE contact. We have recruited a PPIE group who have experienced unexpected bleeding while using hormone replacement therapy (HRT), or who have had an urgent referral for suspected womb cancer, whether or not they were using HRT. Our first meeting focussed on the research questions and study design. The second and third meetings will focus on discussing the results and how best to share them.

### Objectives

1. To describe the incidence of unscheduled bleeding, by HRT use.
2. To describe the incidence of urgent referrals for suspected endometrial cancer, by HRT use.
3. To describe the incidence of endometrial cancer in women referred for suspected endometrial cancer, by HRT use.
4. Diagnostic accuracy of unscheduled bleeding for endometrial cancer diagnosis within one year

### Study design

The ORCHID-e database contains N=34,280,445 anonymised EHR from patients in 1,186 GP practices across England. The GP practices were recruited to be approximately representative across regions, ethnicity, socio-economic status, and rurality. We will utilise a matched cohort design using EHR in the ORCHID-e database. Women starting HRT for postmenopausal symptoms will be matched to women of a similar age who have not started HRT.

### Feasibility counts

The ORCHID-e database contains approximately 400,000 women aged over 40 years who are new users of HRT, and 343 of these were later diagnosed with endometrial cancer. In another study that we did (unpublished) we estimated that about 50% of women experiencing PMB received an urgent referral for suspected cancer. A recent meta-analysis, found that about 9% of women referred for post-menopausal bleeding had endometrial cancer [8].

Based on these statistics and the 343 diagnoses of endometrial cancer we would expect about 3811 urgent referrals and 7622 consultations for PMB within the HRT new user group in our study. These numbers should be enough to calculate proportions with reasonable numbers in the numerator (>=5) for one-way and two-way groupings e.g. proportions of women with referral for suspected endometrial cancer by age and calendar year.

### Study population

All women of age ≥40 years and with time in the data within the study period (January 2015- March 2025) will be eligible for the study. They will become eligible to enter the study at the latest of reaching age 40 years and 1 January 2015 and one year after the registration date at their current practice. Women with a prescription for a HRT oestrogen-containing product in the one-year before the eligibility date will be excluded from the study. Women will no longer be eligible for the study after a record for hysterectomy or a diagnosis code for endometrial cancer.

#### Inclusion criteria

- At least one year registered at the current GP practice whilst aged ≥40 years during the study period.
- No prescription for oestrogen containing HRT in the one-year before becoming eligible for the study

#### Exclusion criteria

- Continuing or past use of HRT as defined by at least one oestrogen containing HRT prescription in the year before becoming eligible for the study.
- Diagnosis of endometrial cancer on or before index date
- Hysterectomy on or before index date. Although some women who have had a partial hysterectomy may receive HRT, we have not found it possible to distinguish between types of hysterectomy using codes recorded in Primary Care.

### Exposure

The exposure will be initiation of HRT for menopausal symptoms, involving both oestrogen and progestogen. Progestogen therapy is often given via an intrauterine system (IUS), but these may not be well recorded in primary care (some IUS devices are placed in sexual health clinics). Therefore, a prescription for progestogen therapy or IUS procedure will not be a requirement for entry into the study population. We will define exposure as a single prescription for a HRT oestrogen-containing product on or after the date of becoming eligible for the study. Index date is the date of this first prescription. Each women in the exposed group will be matched on age at index date to up to four women who have not received a prescription for a HRT-containing oestrogen product on or prior to this date. Index date for the unexposed will be the index date of the matched exposed woman.

We will also extract information on progestogen containing prescriptions on or before the index date. We will use this information for describing the population and for subgroup and sensitivity analysis. Women with a prescription for an oestrogen containing product in the year before they are eligible to enter the study will classified as continuing or past users of HRT and will be excluded from the study.

### Covariates

Covariates, including the following variables will be extracted for the period before the index date. They will be used for describing the study population, stratified analyses and adjusting for confounding.

- Characteristics of prescriptions for products containing progestogens on or before the index date (IUS up to 5 years before index), other progestogen up to 90 days before index.
- Characteristics of index HRT prescription e.g. oral/transdermal, sequential/combined, different types of oestrogens.
- Age at index
- Date of birth
- Calendar year of index date
- Index of multiple deprivation (IMD)
- Urban or rural residence (U/R)
- Ethnicity
- Geographical region
- Smoking
- Menopause code
- Diagnosis of ovarian cancer or breast cancer or cervical cancer
- Unscheduled bleeding or urgent referral for suspected gynaecological cancer
- Adenomyosis
- Risk factors for endometrial cancer if available [11]:
  ○ Polycystic ovarian syndrome
  ○ Endometrial hyperplasia
  ○ Endometriosis
  ○ Ever use of oral contraceptives (possibly protective)
  ○ BMI
  ○ Diabetes
  ○ Family history/genetic predisposition e.g. Lynch syndrome

### Follow-up

Women in the study population will be followed up until the earliest of change of exposure status, study end, death/deregistering, hysterectomy or diagnosis of endometrial cancer. Stopping HRT will initially be defined as a gap of more than 90 days between prescriptions for oestrogen containing HRT, as done by another study [12]. For those in the unexposed group who go on to start HRT, follow-up will be censored at the date of the first prescription. However these women will re-enter the study in the exposed group and will be matched to unexposed who are eligible and unexposed at the new index date.

### Outcomes

- Unscheduled bleeding: This will be defined by a code for postmenopausal bleeding.
- Urgent referral for suspected endometrial cancer. This will be defined as an urgent referral for suspected gynaecological cancer made within 7 days of an unscheduled bleeding code.
- Diagnosis of endometrial cancer within one year of unscheduled bleeding/urgent referral code.

### Statistical analysis

#### Description of the study population

- Baseline characteristics for unexposed and exposed groups including sociodemographic characteristics and risk factors for endometrial cancer.
- Characteristics of index HRT prescriptions
- Characteristics of HRT progestogen prescriptions on or before index
- Characteristics of follow-up/HRT prescribing e.g. total number of months of HRT, changes in HRT prescription where this can be tracked.

#### 1) To describe the incidence of unscheduled bleeding by HRT use

- Incidence proportions for unscheduled bleeding episodes for HRT users and non-users. Incidence will be calculated by 6-month categories of time since index.
- Adjusted hazard ratios for unscheduled bleeding in different periods after index for exposed vs unexposed.
- Analyses will be done overall and by subgroups of age, calendar year, sociodemographic variables, risk factors for endometrial cancer, and types of HRT at index (e.g. oral/transdermal, sequential/combined, different types of oestrogens and progestogens).

#### 2) To describe the incidence of urgent referrals for suspected endometrial cancer by HRT use

- Incidence proportions for urgent referrals for endometrial cancer for HRT users and non- users
- Numbers of women with more than one referral
- Adjusted hazard ratios for urgent referrals in different periods after index for exposed vs unexposed.
- Analyses will be done overall and by 6-month categories of time since index, age, calendar year, sociodemographic variables, risk factors for endometrial cancer, and types of HRT.

#### 3) To describe the incidence of endometrial cancer in women referred for suspected endometrial cancer by HRT use

- Proportions of urgent referrals, followed by a diagnosis of endometrial cancer in the subsequent year.
- Adjusted hazard ratios for endometrial cancer within one year of referral for exposed vs unexposed.
- Analyses will be done overall and by 6-month categories of time since index, age, calendar year, sociodemographic variables, risk factors for endometrial cancer, and types of HRT.

#### 4) Diagnostic accuracy of unscheduled bleeding for endometrial cancer within one year

- Populations:
  ○ Diagnostic accuracy will be ascertained separately for the women on HRT and those not on HRT
- Index tests:
  ○ Unscheduled bleeding > 6 months after index
  ○ Unscheduled bleeding > 6 months after index plus urgent referral
- Reference test: Diagnosis of endometrial cancer within one year of unscheduled bleeding
- Analysis: incidence of endometrial cancer, sensitivity and specificity, positive predictive value, negative predictive value
- Analyses will be done overall and by subgroups such as time since index, age, calendar year, risk factors for endometrial cancer, types of HRT.

### Secondary and sensitivity analyses

We will conduct sensitivity analyses to check the effect of assumptions made. For example:

- Definitions of stopping HRT (time between prescriptions).
- Time period between a report of unscheduled bleeding and an urgent referral

We will conduct sensitivity analyses in which we restrict the exposed group to HRT users who have prescriptions for progestogen as well as oestrogen, on or before their index date.

If it is possible to describe changes in HRT prescriptions we will describe incidence of unscheduled bleeding in relation to these changes.

### Limitations

- Some women may be taking HRT prescribed by private or secondary care clinics and these prescriptions will not have been recorded in Primary Care.
- The definition of new user of HRT is imperfect.
- We will describe the index prescriptions for HRT; however, dose and type could change over follow-up and it will be hard to fully track these changes. We acknowledge that there may be a higher incidence of unscheduled bleeding in 3 months after prescription change.
- We cannot be certain that our definition of HRT excludes oestrogen only HRT. Inappropriate prescribing of oestrogen only HRT to postmenopausal women is in itself a strong risk factor for endometrial cancer, but we will not be able to detect these prescriptions in this study.
- Orchid-e data is primary care only, so some diagnoses of endometrial cancer could be missed.

## Data Availability

This study is based on an anonymised dataset of electronic health records, and cannot be shared.

## Acknowledgements

We thank our PPIE group including amongst others Jo Lloyd, Dr Sarah Markham, Felicia Bassey for their views, which have contributed to this protocol.

## Ethical approval

ORCHID-E has ethical approval from the Health and Social Care Research Ethics Committee B (HSC REC B), reference number 22/NI/0120 and Section 251 approval from the Health Research Authority (HRA) following advice from the Confidentiality Advisory Group (CAG), reference number 25/CAG/0128.

This research is being conducted as part of the NIHR Cancer Awareness, Screening and Early Detection Policy Research Unit (NIHR206132) and has received approval from the Primary Care Hosted Research Datasets Independent Scientific Committee (PrimDISC), reference number PD- 0031-2023.

## Author contributions

MS, CB and BN conceived the study. MS led in developing the concept and designing the study and wrote the first draft of this manuscript. SD provided input from the clinical perspective. All authors contributed to developing the design of the study. All authors commented on earlier drafts of the protocol. All authors saw and approved the final version.

